# Indirect Genetic Effects on Alcohol Use Disorder and Nicotine Dependence

**DOI:** 10.64898/2026.04.17.26351089

**Authors:** Mannan Luo, Victória Trindade Pons, Michael Zakharin, Jean-Baptiste Pingault, Nathan A. Gillespie, Hanna M. van Loo

## Abstract

Substance use disorders run in families, yet the mechanisms underlying intergenerational transmission remain unclear. We investigated indirect genetic effects, pathways through which parental genotypes influence offspring phenotypes via the family environment, for alcohol use disorder (AUD), nicotine dependence (ND), and related quantitative outcomes, and aimed to identify family environmental factors through which such effects may operate. Using transmitted and non-transmitted polygenic scores (PGS) constructed for problematic alcohol use, tobacco use disorder, and general addiction liability, we analyzed 5972 European-ancestry adult offspring with at least one genotyped parent from the population-based Lifelines cohort (Netherlands). Offspring outcomes included lifetime DSM-5 AUD diagnosis, AUD symptom count, maximum drinks in 24 hours, Fagerström Test for Nicotine Dependence score, and cigarettes per day. AUD findings were meta-analyzed with data from the Brisbane Longitudinal Twin Study (*N* = 1368; Australia). We also examined parent-of-origin effects and mediation by parental substance use and socioeconomic status using structural equation modeling. Transmitted PGS robustly predicted all AUD and ND outcomes (*β* = 0.07-0.16; OR = 1.20 for AUD diagnosis). Non-transmitted PGS, indexing indirect genetic effects, were negligible for all clinical syndrome outcomes. The only significant indirect genetic effect was on cigarettes per day (*β* = 0.03, *p* = 0.01), mediated by parental smoking behavior but not socioeconomic status. These findings indicate that intergenerational transmission of risk for AUD and ND is driven primarily by direct genetic effects, with modest indirect genetic effects on smoking quantity. Larger samples and cross-trait analyses are needed to further elucidate these mechanisms.

## Introduction

Substance use disorders (SUDs), including alcohol use disorder (AUD) and nicotine dependence (ND), run in families [1–3]. Offspring of parents with SUDs have a nearly threefold increased risk of developing the same disorder [4, 5]. However, understanding how this risk transmits across generations remains challenging because parents provide both genes and rearing environments [2]. Many putatively environmental risk factors, such as parental substance use or socioeconomic status, are themselves genetically influenced [6]. This potentially induces gene-environment correlation, making it difficult to disentangle the extent to which intergenerational transmission operates through inherited DNA, the rearing environment, or their interplay [7].

One pathway through which gene-environment correlation arises is indirect genetic effects, whereby parental genotypes influence offspring outcomes through the environments parents provide [8, 9]. Although often described as genetic nurture (e.g., parenting processes within the family), indirect genetic effects can also shape offspring outcomes through broader pathways that do not directly involve within-family nurturing [10]. For example, parental genetic liability to AUD may contribute to heavier drinking that normalizes alcohol use in the home [11], or to functional impairment that reduces socioeconomic status and affects neighborhood selection [12], both of which could increase offspring AUD risk independently of parentally transmitted alleles. The transmitted/non-transmitted polygenic score (PGS) design leverages parent-offspring genetic data to separate these pathways by partitioning parental alleles into (i) transmitted PGS (PGS_T_), capturing the combined influence of direct and indirect genetic effects, and (ii) non-transmitted PGS (PGS_NT_), indexing indirect genetic effects independently of direct transmission [8, 9].

Previous studies, including our own, have detected modest indirect genetic effects on quantitative substance use outcomes such as cigarettes per day [13–15]. However, it remains unclear whether these effects generalize to clinically defined SUDs in the general population, specifically AUD and ND, two of the most prevalent and burdensome SUDs worldwide [16, 17]. To date, only one study (*N* = 4,806) has reported indirect genetic effects on AUD, in a high-risk sample enriched for familial alcohol problems [18]. It is also unknown whether such effects differ by parent-of-origin in clinical outcomes, as aggregating maternal and paternal PGS could obscure parent-specific direct [19] and indirect genetic effects [8]. Furthermore, parental substance use has been identified as a plausible mediating pathway for indirect genetic effects on offspring non-clinical outcomes such as smoking [13, 14], yet the mechanisms underlying such effects on clinical SUDs remain poorly understood. In particular, it is unclear whether they operate through parental substance use [20, 21] or socioeconomic disadvantage [22, 23], a transdiagnostic risk factor for SUDs.

To address these gaps, we constructed PGS_T_ and PGS_NT_ to investigate direct and indirect genetic effects on offspring AUD, ND, and related quantitative phenotypes. Primary analyses were conducted in the population-based Lifelines cohort (the Netherlands). Given that indirect genetic effects are typically small [13, 14, 24, 25], we additionally meta-analyzed Lifelines with the Brisbane Longitudinal Twin Study (BLTS, Australia) for AUD outcomes, the only phenotype with comparable diagnostic assessments across cohorts, to increase power and assess cross-population generalizability. This study aimed to: (i) examine whether parental genotypes show indirect genetic effects (indexed by PGS_NT_) on AUD and ND, using both substance-specific PGS and a general addiction liability PGS; (ii) evaluate whether maternal and paternal indirect genetic effects differ in magnitude; and (iii) elucidate whether these effects are mediated by parental substance use, socioeconomic status (parental education and household income), or both.

## Materials and methods

The study protocol was preregistered with the Open Science Framework (https://osf.io/xu9w7/); deviations are described in the **Supplementary Methods.**

### Participants

Primary analyses were conducted in the Lifelines cohort. Lifelines is a multi-disciplinary prospective population-based cohort study examining in a unique three-generation design, the health and health-related behaviors of 167729 persons living in the North of the Netherlands. It employs a broad range of investigative procedures in assessing the biomedical, socio-demographic, behavioral, physical and psychological factors which contribute to the health and disease of the general population, with a special focus on multi-morbidity and complex genetics. Data collection was accomplished across three assessments waves: the baseline (2007-2013), wave 2 (2014-2017), and wave 3 (2019-2023). Detailed information on the study design and sample characteristics has been described elsewhere [26, 27]. For the present study, we included 19233 genotyped adult offspring with at least one genotyped parent, comprising 15966 parent-offspring pairs (father-child or mother-child) and 3267 complete trios (mother-father-offspring). Analytic sample sizes varied by outcome availability, ranging from 2083 to 5972.

For the AUD meta-analysis, we additionally included 1368 adult offspring with at least one genotyped parent from the BLTS [28], yielding a combined meta-analytic sample of 6229 offspring (Lifelines: *n* = 4861; BLTS: *n* = 1368). All other analyses, including ND-related outcomes, parent-of-origin effects, and mediation analyses, were conducted in Lifelines only due to limited data availability in BLTS.

### Phenotypic Measures

#### Alcohol use disorder

In Lifelines, lifetime AUD was assessed at wave 3 (2019-2023) using DSM-5 criteria among ever-drinkers who reported retrospectively on their period of heaviest use. Lifetime AUD diagnosis was defined as endorsement of at least two DSM-5 symptoms [29]. To handle missing items while preserving diagnostic certainty, we applied a conservative PhenX Toolkit algorithm (**Supplementary Methods**). We also examined two continuous AUD-related outcomes to capture dimensional variation and improve statistical power: AUD symptom count (AUDsx) and maximum alcoholic drinks consumed in 24 hours (MaxDrinks), a proxy for binge drinking propensity [18]. In BLTS, lifetime AUD was assessed via self-report questionnaire using the same DSM-5 criteria (**Supplementary Methods**).

#### Nicotine dependence

Lifetime nicotine dependence was assessed at wave 3 using the Fagerström Test for Nicotine Dependence (FTND) [30] in ever-smokers meeting a lifetime exposure threshold for at least one product (≥100 cigarettes or e-cigarettes, ≥25 cigars, ≥50 cigarillos, or ≥40 pipe sessions). Participants reported retrospectively on their period of heaviest tobacco use (**Supplementary Methods**). Those not meeting any exposure threshold were not administered the FTND questionnaire. FTND scores range from 0 to 10, with higher scores indicating greater dependence. We included lifetime average cigarettes per day (CPD) assessed at baseline, as a complementary severity indicator that correlates strongly with FTND [31] and has been shown to index genetic liability to dependence [32].

#### Parental socioeconomic status

Parental socioeconomic status was indexed by years of education and equivalized household income, assessed via self-report at baseline (2006–2013). Educational attainment was derived from participants’ highest obtained educational degree, converted to the corresponding number of years of education required to complete that degree, following previous research in Lifelines [33]. Monthly household income was equivalized by dividing the midpoint of each income category by the square root of household size, thereby accounting for differences in household composition. For interpretability, income was rescaled by dividing by 100, so that estimates reflect differences per 100-euro change in equivalized monthly household income at baseline.

### Genotyping and polygenic scores

Details on genotyping, quality control, imputation, inference of transmitted and non-transmitted alleles, and PGS construction are provided in the **Supplementary Methods**. Briefly, 79988 Lifelines participants were genotyped across three batches and imputed using the Haplotype Reference Consortium v1 panel. Analyses were restricted to participants of European ancestry to minimize population stratification.

We used Haplotype-based Inference of Non-Transmitted Alleles (HINTA) [33], a validated method that overcomes a key limitation of standard transmitted/non-transmitted PGS designs: the requirement for complete parent-offspring trios. By inferring non-transmitted alleles from haplotype comparisons and recombination patterns, HINTA can reliably reconstruct parental non-transmitted alleles even when only one parent is genotyped, achieving 99.8% concordance with standard trio-based pseudo-control phasing approach [34]. This enables accurate distinction of transmitted and non-transmitted alleles in both parent-offspring duos and complete trios, increasing sample size, statistical power, and the generalizability of indirect genetic effect estimates.

Transmitted (PGS_T_) and non-transmitted (PGS_NT_) polygenic scores were constructed using summary statistics from the largest independent genome-wide association studies (GWAS) of problematic alcohol use (PAU) [35], tobacco use disorder (TUD) [36], and a general addiction factor for substance use disorders (SUDs) derived from a multivariate GWAS of problematic alcohol use, problematic tobacco use, cannabis use disorder, and opioid use disorder [37]. Variant weights were estimated using LDpred2-auto [38] and applied in PLINK 1.9 [39] to construct PGSs based on the transmitted and non-transmitted allele datasets.

To maximize statistical power for estimating overall effects, maternal and paternal transmitted and non-transmitted PGS were summed into combined parental scores. The missing parental PGS_NT_ values among parent-offspring pairs were imputed using the mean of observed parents, following previous work [8, 33]. Parent-of-origin and mediation analyses used separate non-imputed maternal and paternal scores (PGS_T_m_, PGS_NT_m,_ PGS_T_f_, PGS_NT_f_), with missingness handled by full-information maximum likelihood (FIML) in *lavaan* [40]. All PGSs were residualized on the first ten genetic principal components and standardized within each genotyping array to control for population stratification and batch effects.

### Statistical analysis

All analyses were conducted in R version 4.2.1 [41].

#### Overall indirect genetic effects on AUD and ND

We used mixed-effects regression models to examine associations between parental PGS_T_ and PGS_NT_ with five offspring outcomes, including AUD diagnosis, AUDsx, MaxDrinks, FTND sum score, and CPD. Continuous outcomes were analyzed using mixed-effects linear regression with *lmerTest* package [42], and dichotomous outcomes using mixed-effects logistic regression with *GLMMadaptive* [43]. Models were specified as: *Outcome_i_* = *β*_1_*PGS_Ti_* + *β*_2_*PGS_NTi_* + *β*_3_*Sex_i_* + *β*_4_*Age_i_* + 1*|Family/D* + *e_i_*. Family ID was included as a random effect (intercept) to account for sibling relatedness, with offspring sex and age as covariates. Consistent with Kong et al. [8], the direct genetic effect (*β*_DGE_) was estimated as the difference between the transmitted and non-transmitted PGS effects (i.e., *β*_T_ *− β*_NT_). Because PGS_T_ captures both direct and indirect genetic effects, subtracting β_NT_ isolates the direct genetic transmission component. To correct for multiple testing across the five correlated outcomes, the effective number of independent tests was estimated using Galwey’s method (*M*_eff_ = 4) with *poolr* [44], and a Šidák-corrected significance threshold of *p* < .013 was applied to balance family-wise error rate control with statistical power [45].

#### Meta-analysis on AUD

Given the modest sample size for clinical AUD in Lifelines and small effect sizes typically expected for indirect genetic effects, we meta-analyzed transmitted and non-transmitted PGS effects on AUD across Lifelines and BLTS. Regression models were fitted separately in each cohort using identical model specifications, as described above. Cohort-specific effect estimates and standard errors were combined using fixed-effects inverse-variance-weighted meta-analysis with *metafor* [46].

#### Parent-of-Origin effects

Unlike the mixed-effects regressions above, which combined maternal and paternal PGS to maximize power, we next fitted structural equation models (SEMs) in *lavaan* [47] to estimate all four PGS components simultaneously (PGS_T_m_, PGS_NT_m_, PGS_T_f_, PGS_NT_f_). This enabled formal testing of differences between maternal and paternal effects (both transmitted and non-transmitted) using likelihood ratio tests (*lavTestLRT*), comparing models in which maternal and paternal paths were constrained to equality against unconstrained models. Models were estimated using FIML, incorporating all available data without listwise deletion or prior imputation [40]. Offspring sex and age were included as covariates, and family ID was specified as a clustering variable with robust standard errors to account for sibling relatedness. All four PGS components were modeled simultaneously with freely estimated covariances, partially accounting for correlations between maternal and paternal PGS induced by assortative mating (non-random mate selection based on phenotypic similarity), and thereby reducing upward bias in indirect genetic effect estimates, although residual bias from multigenerational assortative mating cannot be fully excluded [48, 49].

#### Mediation analyses

To identify which environmental pathways mediated indirect genetic effects, we conducted SEM-based mediation analyses in *lavaan*, restricted to outcomes with significant indirect genetic effects. Parental substance use, parental education, and household income were examined as candidate mediators. Each mediator was initially evaluated in a separate single-mediator model to assess its contribution independently. Significant mediators were then entered into a joint parallel multiple-mediator model to estimate their unique contributions after mutual adjustment. Maternal and paternal pathways were modeled simultaneously to enable parent-specific comparisons, and assortative mating was partially accommodated as described above. Confidence intervals for mediation effects and maternal-paternal difference parameters were obtained using Monte Carlo simulation with *semTools* (2,000 replications) [50]. Statistical significance was inferred when the 95% confidence intervals excluded zero.

## Results

In Lifelines, the analytic sample comprised 5972 adult offspring (mean age = 42.5 years, SD = 10.4), of whom 64.1% were female. Among ever-smokers meeting a lifetime exposure threshold for at least one tobacco product (*n* = 2083), the mean FTND score was 2.11 (SD = 2.16), reflecting nicotine dependence severity during their period of heaviest use. Among all ever-smokers, mean lifetime average CPD was 10.24 (SD = 6.00). Among ever-drinkers, the mean AUD symptom count was 0.88 (SD = 1.31), and 20.6% met criteria for lifetime AUD diagnosis (≥2 symptoms). Sample characteristics did not differ substantially between parent-offspring pairs and complete trios (**Table 1**). Bivariate correlations among PGSs and offspring outcomes are reported in the **Supplementary Tables 1 and 2**.

**Table 1.** Descriptive characteristics of Lifelines participants.

|  | **All** |  | **Pairs** |  | **Trios** |  | ***p*** |
| --- | --- | --- | --- | --- | --- | --- | --- |
|  | ***N*** | ***M*±*SD* or *n* (%)** | ***N*** | ***M*±*SD* or *n* (%)** | ***N*** | ***M*±*SD* or *n* (%)** |  |
| Age (years) at assessment | 5989 | 42.48 ± 10.44 | 4956 | 42.93 ± 10.36 | 1033 | 40.33 ± 10.53 | <.001 |
| Sex, female | 5989 | 3838 (64.1%) | 4956 | 3206 (64.7%) | 1033 | 632 (61.2%) | .036 |
| **Alcohol use disorder** |  |  |  |  |  |  |  |
| AUD diagnosis, yes | 4861 | 1002 (20.61%) | 4009 | 827 (20.6%) | 852 | 175 (20.5%) | .991 |
| AUDsx | 4862 | 0.88 ± 1.31 | 4009 | 0.88 ± 1.31 | 853 | 0.88 ± 1.28 | .979 |
| MaxDrinks | 4762 | 12.67±7.47 | 3926 | 12.61 ± 7.43 | 836 | 12.95 ± 7.61 | .237 |
| **Nicotine dependence** |  |  |  |  |  |  |  |
| FTND sum score | 2083 | 2.11 ± 2.16 | 1762 | 2.15 ± 2.15 | 321 | 1.90 ± 2.19 | .058 |
| CPD* | 5972 | 10.24 ± 6.00 | 5134 | 10.34 ± 6.06 | 838 | 9.59 ± 5.57 | <.01 |

Note. *N* = Sample size, *M* = Mean, *SD* = Standard deviation. Chi-square test for binary outcomes and t-test for continuous outcomes when comparing the pairs versus trios on AUD- and ND-related outcomes. AUDsx =DSM-5 alcohol use disorder symptom count; MaxDrinks = maximum number of alcoholic drinks consumed in 24 hours; FTND = Fagerström Test for Nicotine Dependence; CPD = cigarettes per day; \*CPD was assessed at baseline. Sex assigned at birth was self-reported at baseline enrollment in Lifelines and used in all analyses.

### Indirect genetic effects on AUD and ND

We examined associations of transmitted (PGS_T_) and non-transmitted (PGS_NT_) polygenic scores with offspring AUD- and ND-related outcomes, including clinical outcomes (lifetime AUD diagnosis and AUD symptom count) and quantitative measures (MaxDrinks, FTND sum score, and CPD) (**Table 2**).

**Table 2.** Associations of transmitted (PGS_T_) and non-transmitted (PGS_NT_) polygenic scores with offspring alcohol use disorder (AUD) and nicotine dependence (ND) outcomes in Lifelines.

| Predictors | Outcomes | N | PGS<sub>T</sub> |  |  | PGS<sub>NT</sub> |  |  | DGE |
| --- | --- | --- | --- | --- | --- | --- | --- | --- | --- |
| | | | $\beta_T$ (SE)/OR<sub>T</sub> | 95% CI | p | $\beta_{NT}$ (SE)/OR<sub>NT</sub> | 95% CI | p | $\beta_{DGE}$ |
| Substance-specific PGS |  |  |  |  |  |  |  |  |  |
| PGS<sub>PAU</sub> | AUD diagnosis | 4861 | 1.196 | 1.105, 1.294 | <.001 | 0.959 | .888, 1.035 | .277 | — |
|  | AUD<sub>sx</sub> | 4862 | .078 (.014) | .050, .105 | <.001 | -.007 (.014) | -.035, .020 | .598 | .071 |
|  | MaxDrinks | 4762 | .068 (.012) | .044, .092 | <.001 | -.004 (.012) | -.028, .020 | .721 | .064 |
| PGS<sub>TUD</sub> | FTND<sub>sum</sub> | 2083 | .161 (.022) | .118, .204 | <.001 | .031 (.022) | -.013, .074 | .168 | .130 |
|  | CPD | 5972 | .127 (.013) | .101, .152 | <.001 | .033 (.013) | .007, .059 | .012 | .094 |
| General addiction factor |  |  |  |  |  |  |  |  |  |
| PGS<sub>SUD</sub> | AUD diagnosis | 4861 | 1.113 | 1.031, 1.202 | .006 | .975 | .903, 1.053 | .520 | — |
|  | AUD<sub>sx</sub> | 4862 | .061 (.014) | .033, .089 | <.001 | -.014 (.014) | -.042, .014 | .322 | .047 |
|  | MaxDrinks | 4762 | .068 (.012) | .044, .092 | <.001 | -.018 (.012) | -.042, .006 | .140 | .050 |
|  | FTND<sub>sum</sub> | 2083 | .121 (.022) | .077, .165 | <.001 | .0002 (.022) | -.043, .043 | .990 | .121 |
|  | CPD | 5972 | .116 (.013) | .091, .141 | <.001 | .021 (.013) | -.004, .046 | .111 | .095 |

Note. Mixed-effects regression models included up to 5,972 adult offspring with at least one genotyped parent and available outcome data. Family ID was included as a random intercept to account for sibling relatedness, with offspring sex and age as covariates. Continuous outcomes were analyzed using linear mixed-effects models and lifetime AUD diagnosis using mixed-effects logistic regression. PAU = problematic alcohol use; TUD = tobacco use disorder; SUD = substance use disorders; AUD<sub>sx</sub> = AUD symptom count; AUD<sub>sx</sub> = DSM-5 alcohol use disorder symptom count; MaxDrinks = maximum number of alcoholic drinks consumed in 24 hours; FTND = Fagerström Test for Nicotine Dependence; CPD = cigarettes per day; DGE = direct genetic effect. $\beta$ = standardized regression coefficient; OR = odds ratio; SE = standard error; CI = 95% confidence interval. *p*-values are unadjusted. Šidák-corrected significance threshold: $p < .013$ . $\beta_{DGE} = \beta_T - \beta_{NT}$ for continuous outcomes.

Transmitted PGS for both substance-specific (PGS _T_PAU_, PGS_T_TUD_) and general addiction PGS (PGS_T_SUD_) showed robust associations with corresponding outcomes (*β*_T_ = .061-.161 for continuous traits; OR = 1.113-1.196 for AUD diagnosis; all *p* < .001). In contrast, indirect genetic effects, indexed by PGS_NT_, were largely absent. Of all associations tested, only one survived multiple-testing correction: parental PGS_NT_TUD_ was modestly associated with offspring CPD (*β*_NT_ = .033, *p* = .012). No other significant indirect genetic effects were observed for AUD diagnosis, AUDsx, MaxDrinks, or FTND sum score.

Direct genetic effects were quantified as *β*_DGE_ = *β*_T_ − *β*_NT_, following Kong et al. [8]. These effects were most pronounced for FTND sum score (*β*_DGE_ = .130) and CPD (*β*_DGE_ = .094) using PGS_TUD_. The sole significant indirect genetic effect, parental PGS_NT_TUD_ on offspring CPD, was approximately 35% of the corresponding direct genetic effect.

### Meta-analysis of AUD

To enhance statistical power and assess cross-population generalizability, PGS_T_ and PGS_NT_ effects on AUD were meta-analyzed across Lifelines and BLTS (total *N* = 6229; **Supplementary Table 3 and Figure 1**). Indirect genetic effects remained nonsignificant for both PGS_NT_PAU_ and PGS_NT_SUD_, consistent with Lifelines findings.

### Parent-of-origin effects

To examine whether maternal and paternal indirect genetic effects differed, transmitted and non-transmitted PGS were decomposed by parent-of-origin (**Table 3**). Both maternal and paternal PGS_T_ significantly predicted all offspring outcomes, with comparable effect sizes across parents (e.g., PGS _T_TUD_ on FTND: β_maternal_ =.104; β_paternal_ = .119). For indirect genetic effects, only paternal PGS_NT_TUD_ showed a nominal association with offspring CPD (*β* = .046, *p* = .017), which did not survive correction for multiple testing. No parent-specific indirect genetic effects were observed for ND or AUD outcomes using PGS_NT_SUD_. Likelihood ratio tests indicated no significant maternal–paternal differences in either transmitted or non-transmitted PGS effects for any outcome (all *p* > .25).

**Table 3.** Parent-of-Origin effects on alcohol use disorder and nicotine dependence in Lifelines: transmitted and non-transmitted polygenic scores split by paternal and maternal haplotypes.

|  | Maternal PGS<sub>T</sub> |  |  | Maternal PGS<sub>NT</sub> |  |  | Paternal PGS<sub>T</sub> |  |  | Paternal PGS<sub>NT</sub> |  |  |
| --- | --- | --- | --- | --- | --- | --- | --- | --- | --- | --- | --- | --- |
| | $\beta$ (SE) | 95% CI | *p* | $\beta$ (SE) | 95% CI | *p* | $\beta$ (SE) | 95% CI | *p* | $\beta$ (SE) | 95% CI | *p* |
| **Substance-specific PGS** |  |  |  |  |  |  |  |  |  |  |  |  |
| *PGS<sub>PAU</sub>* |  |  |  |  |  |  |  |  |  |  |  |  |
| AUDsx | .047 (.014) | .019, .076 | .001 | -.008 (.016) | -.041, .024 | .618 | .063 (.014) | .036, .089 | <.001 | -.007 (.021) | -.047, .033 | .735 |
| MaxDrinks | .044 (.012) | .021, .068 | <.001 | -.001 (.014) | -.028, .026 | .929 | .051 (.012) | .027, .074 | <.001 | -.017 (.018) | -.053, .019 | .360 |
| *PGS<sub>TUD</sub>* |  |  |  |  |  |  |  |  |  |  |  |  |
| FTND<sub>sum</sub> | .104 (.022) | .062, .147 | <.001 | .007 (.026) | -.043, .058 | .776 | .119 (.022) | .077, .162 | <.001 | .053 (.034) | -.013, .119 | .117 |
| CPD | .078 (.013) | .053, .103 | <.001 | .016 (.016) | -.016, .047 | .333 | .095 (.013) | .070, .120 | <.001 | .046 (.019) | .008, .084 | .017 |
| **General addiction factor** |  |  |  |  |  |  |  |  |  |  |  |  |
| *PGS<sub>SUD</sub>* |  |  |  |  |  |  |  |  |  |  |  |  |
| AUDsx | .053 (.019) | .016, .091 | .005 | -.035 (.023) | -.081, .011 | .142 | .057 (.017) | .025, .090 | .001 | .007 (.028) | -.049, .062 | .809 |
| MaxDrinks | .055 (.012) | .032, .079 | <.001 | -.018 (.014) | -.045, .010 | .203 | .041 (.012) | .017, .065 | .001 | -.016 (.018) | -.052, .020 | .378 |
| FTND<sub>sum</sub> | .093 (.022) | .050, .135 | <.001 | -.025 (.025) | -.074, .023 | .308 | .078 (.023) | .032, .123 | .001 | .044 (.034) | -.024, .111 | .204 |
| CPD | .077 (.013) | .051, .103 | <.001 | .028 (.016) | -.002, .059 | .071 | .081 (.012) | .057, .105 | <.001 | .003 (.020) | -.036, .042 | .879 |

Note. Structural equation models (SEMs) estimated maternal and paternal transmitted (PGS<sub>T</sub>) and non-transmitted (PGS<sub>NT</sub>) effects. Models were fitted in *lavaan* using full information maximum likelihood (FIML) with all available data from the genotyped family sample (parents and offspring; *N* = 19,233), with outcomes measured in adult offspring only; analytic sample sizes per outcome match Table 2.
Standardized coefficients ( $\beta$ ), standard errors (SE), 95% confidence intervals (CIs), and unadjusted *p* values are reported. All models adjusted for offspring sex and age, and accounted for sibling relatedness using family ID as a clustering variable with robust standard errors. Because FIML for binary outcomes (AUD diagnosis) is not supported in *lavaan*, analyses were restricted to the four continuous outcomes. The Šidák-corrected significance threshold was *p* < .013.

### Mediation analysis

Given that significant indirect genetic effects were observed only for CPD, we examined whether these effects operated through parental smoking, parental education, and household income (**Supplementary Table 4**). In single-mediator models, parental smoking emerged as the primary mediator, with maternal PGS_NT_TUD_ showing stronger mediation via maternal smoking (*β* = .014, 95% MC CI [.007, .021]) and paternal PGS_NT_TUD_ via paternal smoking (*β* = .007 [.002, .011]). Significant mediation was also observed for transmitted PGS_T_TUD_ via maternal smoking (*β* = .022 [.015, .030]) and paternal smoking (*β* = .004 [.001, .009]). Maternal education showed weaker evidence of mediation for PGS_NT_TUD_ (*β* = .004 [.001, .007]) but no significant mediation via paternal education. Household income did not mediate effects for either parent.

To assess whether maternal education represented an independent mechanism or was confounded by parental smoking, we fitted a parallel multiple-mediator model including both mediators simultaneously (**Figure 1**). Parental smoking estimates were unchanged, whereas mediation via parental education was attenuated to non-significance after adjustment for smoking. The transmitted mediation effects (PGS_T_TUD_) via maternal smoking were significantly larger than the corresponding paternal effects (*β*_diff_ = .018 [.009, .027]); no other maternal-paternal differences were significant.

**Figure 1.**
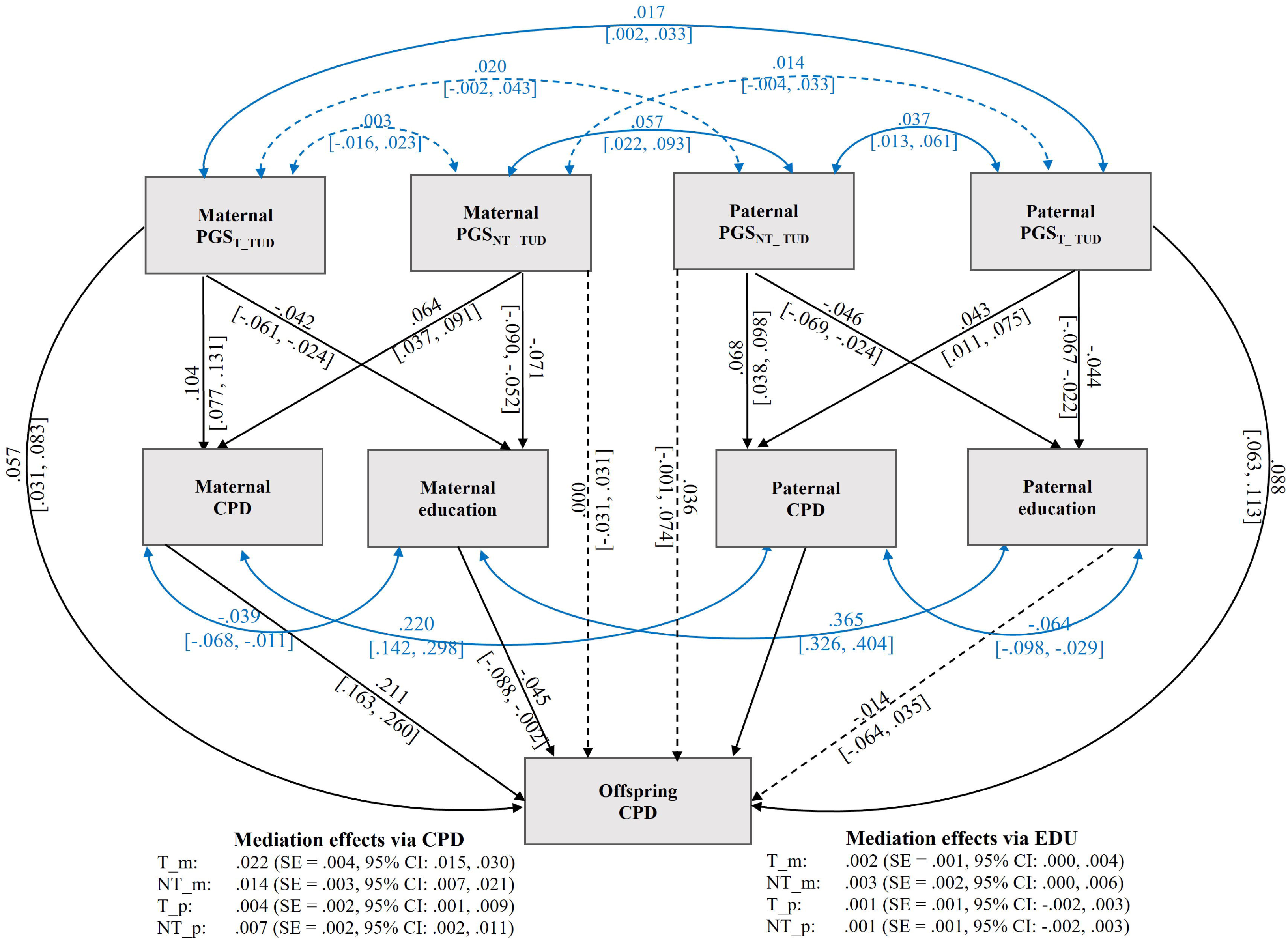
Parallel mediation model of offspring smoking quantity via parental smoking and parental education. *Note.* Structural equation model testing maternal and paternal smoking quantity (CPD) and parental education as parallel mediators of associations between (PGS_T_TUD_) and non-transmitted (PGS_NT_TUD_) polygenic scores and offspring CPD. Models were estimated using full information maximum likelihood (FIML) with cluster-robust standard errors, adjusting for offspring sex and age. The total analytic sample passed to the model was *N* = 19,233. Observed Ns varied across variables due to data availability: PGS_T_ = 19,233 (both parents); PGS_NT_ = 13,236 (maternal) and 9,264 (paternal); parental CPD *N* = 6,472 (maternal) and 5,152 (paternal); parental education *N* = 12,965 (maternal) and 9,095 (paternal); offspring CPD *N* = 5,972. Solid paths indicate associations with 95% Monte Carlo confidence intervals excluding zero; dashed paths indicate nonsignificant associations. Double-headed arrows denote freely estimated covariances between maternal and paternal PGS and parental phenotypes, partially accounting for assortative mating between parents. All path coefficients are standardized.

## Discussion

This study provides the first comprehensive population-based examination of indirect genetic effects on AUD and ND. Using transmitted and non-transmitted polygenic scores in 5972 parent-offspring pairs and trios, supplemented by a meta-analysis of AUD across cohorts (total *N* = 6229), we found that intergenerational transmission of AUD and ND occurs primarily through direct genetic effects. Indirect genetic effects were largely absent, with the exception of offspring smoking quantity (CPD). Mediation analyses suggested parental smoking as a plausible environmentally-mediated pathway, with minimal contribution from parental socioeconomic factors.

Our null findings for indirect genetic effects on AUD contrast with prior evidence from Thomas et al. [18], who reported such effects on AUD in a high-risk sample enriched for familial alcohol problems. This discrepancy may reflect methodological differences. Thomas and colleagues did not test the total effect of non-transmitted parental PGS on offspring AUD, the standard parameter for indirect genetic effects [8, 9], but instead examined specific mediated pathways (e.g., parental relationship discord). Such mediated effects are difficult to interpret without a significant overall PGS_NT_ effect, as individual pathways can reach significance even when total indirect genetic effects does not [51]. Moreover, these mediator-outcome associations remain observational and susceptible to genetic confounding, so apparent mediation may arise spuriously in the absence of true indirect genetic effects [52]. Finally, their high-risk sample likely captured more extreme environmental adversity, thereby increasing sensitivity to detect indirect genetic effects relative to population-based cohorts.

We observed divergent findings across nicotine outcomes: indirect genetic effects were detectable for lifetime average smoking quantity (CPD) but not for nicotine dependence (FTND). One explanation may be limited statistical power, as the FTND was assessed only at Wave 3 among a subset of ever-smokers meeting stricter inclusion criteria, reducing power to detect small indirect genetic effects relative to the broader CPD sample. Alternatively, this pattern may suggest that CPD and FTND capture distinct dimensions of nicotine-related risk. CPD quantifies cumulative smoking behavior across the lifespan and may be more sensitive to environmental influences such as parental behavioral modeling and household smoking norms [53, 54], whereas FTND captures physiological dependence during the period of heaviest use and may more strongly reflect neurobiological mechanisms and direct genetic liability [32]. Larger studies with more comprehensive smoking phenotypes will be needed to determine whether indirect genetic effects differentially influence behavioral versus physiological dimensions of nicotine dependence

A key contribution of this study is the dissection of intergenerational pathways using joint mediation models that allow parental phenotypes to mediate both transmitted and non-transmitted genetic effects on offspring. Although parental genetic liability for SUDs has been proposed to reduce socioeconomic status and thereby increase offspring substance use risk [13, 55], we found that the indirect genetic effect on offspring CPD operated primarily through parental smoking quantity, while the contribution of parental education was fully attenuated when parental smoking was included in the same model. This is consistent with social learning theory, which posits that transmission may occur through behavioral modeling, whereby children observe and imitate parental behaviors [56]. These findings should be interpreted cautiously, as the pathway from parental phenotype to offspring outcome remains observational and may be confounded by unmeasured environmental factors and residual genetic liability not captured by the PGS [52]. They therefore represent an exploration of plausible environmentally-mediated pathways rather than definitive evidence of causal mechanisms.

This study has several strengths, including its large population-based design, the separation of transmitted and non-transmitted alleles in genotyped parent-offspring pairs and trios, the inclusion of both clinical syndromes and dimensional traits, and the cross-cohort meta-analysis for AUD. Furthermore, SEM-based mediation modeling enabled simultaneous evaluation of multiple environmental pathways while partially accounting for assortative mating. However, several limitations should be noted. First, statistical power was constrained by modest sample sizes for certain outcomes, particularly FTND, and by limited predictive performance of current SUD PGSs [57]. Transmitted PGS effects were modest (*β* = .07–.16), suggesting that these scores capture only a fraction of heritable variance. Together, these factors reduced our ability to detect more nuanced indirect genetic pathways, including parent-of-origin patterns and socioeconomic mediation. Second, parental household income was reported when offspring were already adults, limiting its validity as a proxy for financial resources during critical developmental periods. Third, in addition to genuine indirect genetic effects, PGS_NT_ may capture residual confounding from population stratification and assortative mating [52, 58]. Although we adjusted for population structure using principal components and partially accounted for assortative mating within the SEM, residual bias may remain. Finally, analyses were restricted to individuals of European ancestry, limiting generalizability to diverse populations.

## Conclusion

In this large-scale, population-based study, the intergenerational transmission of risk for AUD and ND appears to be driven primarily by direct genetic effects, with minimal evidence for indirect genetic effects. The exception was a modest effect on offspring smoking quantity, operating through parental smoking rather than broader socioeconomic pathways. However, the absence of detectable indirect genetic effects does not preclude their role in SUDs, as such effects may be subtle and require greater statistical power to detect. Future research combining larger samples, more refined SUD measures, and increasingly predictive polygenic scores will be essential to detect and characterize indirect genetic effects in intergenerational transmission of SUDs.

## Data availability

Individual-level data from the Lifelines Cohort are available under restricted access due to ethical requirements and privacy regulations protecting participant confidentiality. Researchers can apply to use the Lifelines data through the online application system. More information about how to request Lifelines data and the conditions of use can be found on their website (https://www.lifelines.nl/researcher/how-to-apply). GWAS summary statistics used to construct the polygenic scores were obtained from publicly available sources, including alcohol use disorder and substance use disorder GWAS from the Psychiatric Genomics Consortium (https://www.med.unc.edu/pgc/). Tobacco use disorder GWAS summary statistics are available upon request from the corresponding author.

## Acknowledgements

The Lifelines initiative has been made possible by subsidy from the Dutch Ministry of Health, Welfare and Sport, the Dutch Ministry of Economic Affairs, the University Medical Center Groningen (UMCG), Groningen University and the Provinces in the North of the Netherlands (Drenthe, Friesland, Groningen). We acknowledge the services of the Lifelines Cohort Study, the contributing research centers delivering data to Lifelines, and all the study participants. We also thank the twins and their families for their participation in the Brisbane Longitudinal Twin Study.

## Ethical approval and consent to participate

The general Lifelines protocol has been approved by the UMCG Medical ethical committee under number 2007/152. Lifelines uses multiple channels to keep participants engaged and actively involved. For this purpose, Lifelines has a Participant Advisory Board. The Participant Advisory Board is an independent council consisting of active Lifelines participants that represents the common interests of all participants who participate in the Lifelines research. The council does this by providing solicited and unsolicited advice to management on strategic and practical matters that are important to participants. The council meets at least four times a year. The timing and frequency are determined on a demand-driven basis. If the council or Lifelines have relevant decision-making or policy issues, these will be addressed. Lifelines is responsible for providing relevant background information for these issues (such as reports, reports, overviews of research applications).

## References

1. Jacob T, Waterman B, Heath A, True W, Bucholz KK, Haber R, et al. Genetic and environmental effects on offspring alcoholism: new insights using an offspring-of-twins design. Arch Gen Psychiatry. 2003;60(12):1265–72.

2. Kendler KS, Ji J, Edwards AC, Ohlsson H, Sundquist J, Sundquist K. An extended Swedish national adoption study of alcohol use disorder. JAMA Psychiatry. 2015;72(3):211–8.

3. Merikangas KR, Stolar M, Stevens DE, Goulet J, Preisig MA, Fenton B, et al. Familial transmission of substance use disorders. Arch Gen Psychiatry. 1998;55(11):973–9.

4. Uher R, Pavlova B, Radua J, Provenzani U, Najafi S, Fortea L, et al. Transdiagnostic risk of mental disorders in offspring of affected parents: a meta-analysis of family high-risk and registry studies. World Psychiatry. 2023;22(3):433–48.

5. Manhica H, Lundin A, Wennberg P, Danielsson AK. Parental substance use disorders and psychiatric conditions in offspring: A Swedish population-based cohort study with over 1,000,000 individuals. J Psychiatr Res. 2024;179:156–62.

6. Kendler K, Baker J. Genetic influences on measures of the environment: a systematic review. Psychol Med. 2007;37(5):615–26.

7. Jami ES, Hammerschlag AR, Bartels M, Middeldorp CM. Parental characteristics and offspring mental health and related outcomes: a systematic review of genetically informative literature. Transl Psychiatry. 2021;11(1):197.

8. Kong A, Thorleifsson G, Frigge ML, Vilhjalmsson BJ, Young AI, Thorgeirsson TE, et al. The nature of nurture: Effects of parental genotypes. Science. 2018;359(6374):424–8.

9. Bates TC, Maher BS, Medland SE, McAloney K, Wright MJ, Hansell NK, et al. The Nature of Nurture: Using a Virtual-Parent Design to Test Parenting Effects on Children’s Educational Attainment in Genotyped Families. Twin Res Hum Genet. 2018;21(2):73–83.

10. Allegrini AG, Hannigan LJ, Frach L, Barkhuizen W, Baldwin JR, Andreassen OA, et al. Intergenerational transmission of polygenic predisposition for neuropsychiatric traits on emotional and behavioural difficulties in childhood. Nat Commun. 2025;16(1):2674.

11. Meulewaeter F, De Schauwer E, De Pauw SSW, Vanderplasschen W. “I grew up amidst alcohol and drugs:” a qualitative study on the lived experiences of parental substance use among adults who developed substance use disorders themselves. Front Psychiatry. 2022;13:768802.

12. Buu A, Dipiazza C, Wang J, Puttler LI, Fitzgerald HE, Zucker RA. Parent, family, and neighborhood effects on the development of child substance use and other psychopathology from preschool to the start of adulthood. J Stud Alcohol Drugs. 2009;70(4):489–98.

13. Saunders GRB, Liu M, Vrieze S, McGue M, Iacono WG. Mechanisms of parent-child transmission of tobacco and alcohol use with polygenic risk scores: Evidence for a genetic nurture effect. Dev Psychol. 2021;57(5):796–804.

14. Luo M, Trindade Pons V, Gillespie NA, van Loo HM. Genetic nurture in intergenerational transmission of substance use. Nature Communications. 2026.

15. Pasman JA, Smit K, Vollebergh WAM, Nolte IM, Hartman CA, Abdellaoui A, et al. Interplay between genetic risk and the parent environment in adolescence and substance use in young adulthood: A TRAILS study. Dev Psychopathol. 2023;35(1):396–409.

16. MacKillop J, Agabio R, Feldstein Ewing SW, Heilig M, Kelly JF, Leggio L, et al. Hazardous drinking and alcohol use disorders. Nat Rev Dis Primers. 2022;8(1):80.

17. Le Foll B, Piper ME, Fowler CD, Tonstad S, Bierut L, Lu L, et al. Tobacco and nicotine use. Nat Rev Dis Primers. 2022;8(1):19.

18. Thomas NS, Salvatore JE, Kuo SI, Aliev F, McCutcheon VV, Meyers JM, et al. Genetic nurture effects for alcohol use disorder. Mol Psychiatry. 2023;28(2):759–66.

19. Hofmeister RJ, Cavinato T, Karimi R, van der Graaf A, Pajuste F-D, Kronberg J, et al. Parent-of-origin effects on complex traits in up to 236,781 individuals. Nature. 2025.

20. Yule AM, Wilens TE, Martelon M, Rosenthal L, Biederman J. Does exposure to parental substance use disorders increase offspring risk for a substance use disorder? A longitudinal follow-up study into young adulthood. Drug Alcohol Depend. 2018;186:154–8.

21. Latvala A, Kuja-Halkola R, D’Onofrio BM, Jayaram-Lindström N, Larsson H, Lichtenstein P. Association of parental substance misuse with offspring substance misuse and criminality: a genetically informed register-based study. Psychol Med. 2022;52(3):496–505.

22. Lee JO, Cho J, Yoon Y, Bello MS, Khoddam R, Leventhal AM. Developmental pathways from parental socioeconomic status to adolescent substance use: alternative and complementary reinforcement. J Youth Adolesc. 2018;47(2):334–48.

23. Melchior M, Choquet M, Le Strat Y, Hassler C, Gorwood P. Parental alcohol dependence, socioeconomic disadvantage and alcohol and cannabis dependence among young adults in the community. Eur Psychiatry. 2011;26(1):13–7.

24. Wang B, Baldwin JR, Schoeler T, Cheesman R, Barkhuizen W, Dudbridge F, et al. Robust genetic nurture effects on education: A systematic review and meta-analysis based on 38,654 families across 8 cohorts. Am J Hum Genet. 2021;108(9):1780–91.

25. Tubbs JD, Sham PC. Preliminary Evidence for Genetic Nurture in Depression and Neuroticism Through Polygenic Scores. JAMA Psychiatry. 2023;80(8):832–41.

26. Sijtsma A, Rienks J, van der Harst P, Navis G, Rosmalen JGM, Dotinga A. Cohort Profile Update: Lifelines, a three-generation cohort study and biobank. Int J Epidemiol. 2022;51(5):e295–e302.

27. Scholtens S, Smidt N, Swertz MA, Bakker SJ, Dotinga A, Vonk JM, et al. Cohort Profile: LifeLines, a three-generation cohort study and biobank. Int J Epidemiol. 2015;44(4):1172–80.

28. Gillespie NA, Henders AK, Davenport TA, Hermens DF, Wright MJ, Martin NG, et al. The Brisbane Longitudinal Twin Study: Pathways to Cannabis Use, Abuse, and Dependence project-current status, preliminary results, and future directions. Twin Res Hum Genet. 2013;16(1):21–33.

29. Diagnostic and statistical manual of mental disorders: DSM-5™, 5th ed. Arlington, VA, US: American Psychiatric Publishing, Inc.; 2013. xliv, 947–xliv, p.

30. Heatherton TF, Kozlowski LT, Frecker RC, FAGERSTROM KO. The Fagerström test for nicotine dependence: a revision of the Fagerstrom Tolerance Questionnaire. British journal of addiction. 1991;86(9):1119–27.

31. Vink JM, Willemsen G, Beem AL, Boomsma DI. The Fagerström Test for Nicotine Dependence in a Dutch sample of daily smokers and ex-smokers. Addictive Behaviors. 2005;30(3):575–9.

32. Quach BC, Bray MJ, Gaddis NC, Liu M, Palviainen T, Minica CC, et al. Expanding the genetic architecture of nicotine dependence and its shared genetics with multiple traits. Nat Commun. 2020;11(1):5562.

33. Trindade Pons V, Claringbould A, Kamphuis P, Oldehinkel AJ, van Loo HM. Using parent-offspring pairs and trios to estimate indirect genetic effects in education. Genet Epidemiol. 2024;48(4):190–9.

34. Cordell HJ, Barratt BJ, Clayton DG. Case/pseudocontrol analysis in genetic association studies: A unified framework for detection of genotype and haplotype associations, gene-gene and gene-environment interactions, and parent-of-origin effects. Genet Epidemiol. 2004;26(3):167–85.

35. Zhou H, Kember RL, Deak JD, Xu H, Toikumo S, Yuan K, et al. Multi-ancestry study of the genetics of problematic alcohol use in over 1 million individuals. Nat Med. 2023;29(12):3184–92.

36. Toikumo S, Jennings MV, Pham BK, Lee H, Mallard TT, Bianchi SB, et al. Multi-ancestry meta-analysis of tobacco use disorder identifies 461 potential risk genes and reveals associations with multiple health outcomes. Nat Hum Behav. 2024;8(6):1177–93.

37. Hatoum AS, Colbert SMC, Johnson EC, Huggett SB, Deak JD, Pathak G, et al. Multivariate genome-wide association meta-analysis of over 1 million subjects identifies loci underlying multiple substance use disorders. Nat Ment Health. 2023;1(3):210–23.

38. Privé F, Arbel J, Vilhjálmsson BJ. LDpred2: better, faster, stronger. Bioinformatics. 2021;36(22-23):5424–31.

39. Chang CC, Chow CC, Tellier LC, Vattikuti S, Purcell SM, Lee JJ. Second-generation PLINK: rising to the challenge of larger and richer datasets. GigaScience. 2015;4(1).

40. Lee T, Shi D. A comparison of full information maximum likelihood and multiple imputation in structural equation modeling with missing data. Psychol Methods. 2021;26(4):466–85.

41. R Core Team. R: A language and environment for statistical computing: R Foundation for Statistical Computing, Vienna, Austria; 2024.

42. Kuznetsova A, Brockhoff PB, Christensen RHB. lmerTest Package: Tests in Linear Mixed Effects Models. Journal of Statistical Software. 2017;82(13):1 – 26.

43. Rizopoulos D. GLMMadaptive: generalized linear mixed models using adaptive gaussian quadrature. 2023.

44. Galwey NW. A new measure of the effective number of tests, a practical tool for comparing families of non-independent significance tests. Genet Epidemiol. 2009;33(7):559–68.

45. Abdi H. Bonferroni and Šidák corrections for multiple comparisons. Encyclopedia of measurement and statistics. 2007;3(01):2007.

46. Viechtbauer W. Conducting meta-analyses in R with the metafor package. Journal of statistical software. 2010;36:1–48.

47. Rosseel Y. lavaan: an R package for structural equation modeling. Journal of Statistical Software. 2012;48(2):1 – 36.

48. Balbona JV, Kim Y, Keller MC. Estimation of parental effects using polygenic scores. Behav Genet. 2021;51(3):264–78.

49. Young AS. Estimation of indirect genetic effects and heritability under assortative mating. bioRxiv. 2023.

50. Pesigan IJA, Cheung SF. Monte Carlo confidence intervals for the indirect effect with missing data. Behav Res Methods. 2024;56(3):1678–96.

51. Chuong M, Kwong A, Amador C, Haley C, McIntosh A, Adams M. Methodological Considerations When Using Polygenic Scores to Explore Parent-Offspring Genetic Nurturing Effects [version 1; peer review: 2 approved with reservations]. Wellcome Open Research. 2026;11(19).

52. Pingault JB, Allegrini AG, Odigie T, Frach L, Baldwin JR, Rijsdijk F, et al. Research Review: How to interpret associations between polygenic scores, environmental risks, and phenotypes. J Child Psychol Psychiatry. 2022;63(10):1125–39.

53. Mumford EA, Liu W. Growth models of maternal smoking behavior: individual and contextual factors. Subst Use Misuse. 2015;50(10):1261–73.

54. Owusu D, Quinn M, Wang K, Williams F, Mamudu HM. Smokefree home rules and cigarette smoking intensity among smokers in different stages of smoking cessation from 20 low-and-middle income countries. Prev Med. 2020;132:106000.

55. Deak JD, Johnson EC. Genetics of substance use disorders: a review. Psychol Med. 2021;51(13):2189–200.

56. Branje S, Geeraerts S, de Zeeuw EL, Oerlemans AM, Koopman-Verhoeff ME, Schulz S, et al. Intergenerational transmission: Theoretical and methodological issues and an introduction to four Dutch cohorts. Dev Cogn Neurosci. 2020;45:100835.

57. Kember RL, Davis CN, Feuer KL, Kranzler HR. Considerations for the application of polygenic scores to clinical care of individuals with substance use disorders. J Clin Invest. 2024;134(20).

58. Nivard MG, Belsky DW, Harden KP, Baier T, Andreassen OA, Ystrøm E, et al. More than nature and nurture, indirect genetic effects on children’s academic achievement are consequences of dynastic social processes. Nat Hum Behav. 2024;8(4):771–8.

