## Supplemental Information for "Indirect Genetic Effects on Alcohol Use Disorder and Nicotine Dependence"

### Supplementary Methods

The study protocol was preregistered prior to data analysis (OSF: https://osf.io/xu9w7/). There was one deviation from the preregistered plan:

#### Addition of meta-analysis with BLTS

The preregistered protocol specified analyses in the Lifelines cohort only. During the study, we additionally incorporated data from the Brisbane Longitudinal Twin Study (BLTS; *N* = 1,368) to meta-analyze genetic nurture effects on AUD outcomes. This addition was motivated by two considerations. First, genetic nurture effects are expected to be modest in magnitude, and larger samples would be needed to reliably detect such effects for binary outcomes like AUD diagnosis. Second, BLTS provided comparable lifetime AUD measures based on identical DSM-5 criteria and used the same HINTA (Haplotype-based Inference of Non-Transmitted Alleles) approach [1] to infer transmitted and non-transmitted alleles, making it well-suited for cross-cohort synthesis. The meta-analysis was restricted to AUD outcomes (diagnosis and symptom count) due to limited availability of nicotine dependence measures in BLTS. All other analyses, including nicotine dependence outcomes, parent-of-origin effects, and mediation analyses, were conducted in Lifelines as preregistered. This deviation enhances statistical power and enables assessment of cross-cohort generalizability without changing the core hypotheses or analytic framework specified in the preregistration.

#### Measures

Full scoring scripts for FTND and DSM-5 AUD are available on GitHub (<https://github.com/mannanluo/SUD_AUD-FTND_Scoring>).

##### *Nicotine dependence*

Lifetime nicotine dependence in Lifelines was assessed at Wave 3 (2019–2023) using the Fagerström Test for Nicotine Dependence (FTND)[2], a well-validated six-item self-report measure of physical nicotine dependence administered via digital questionnaire. The FTND was administered to participants who reported ever using tobacco and met a minimum lifetime exposure threshold for at least one product: ≥100 cigarettes or e-cigarettes, ≥25 cigars, ≥50 cigarillos, or ≥40 pipe sessions. Participants were asked to report retrospectively on their lifetime period of heaviest

tobacco use. The FTND includes the following six items:

1) how soon after you wake up did you smoke your first cigarette? (0: >60 min, 1: 31-60 min, 2: 6-30 min, 3: within 5 min)
2) did you find it difficult to refrain from smoking in places where it is forbidden? (0: no, 1: yes)
3) which cigarette would you hate most to give up? (0: all others, 1: the first one in the morning)
4) how many cigarettes/ e-cigarettes/ cigars/ cigarillos/ pipes did you usually smoke per day?
5) did you smoke more frequently during the first hours after waking than during the rest of the day? (0: no, 1: yes)
6) did you smoke if you were so ill that you were in bed most of the day? (0: no, 1: yes)

For Item 4, product-specific nicotine scoring thresholds were applied to ensure clinical equivalence across tobacco types: cigarettes/e-cigarettes (0: ≤10; 1: 11–20; 2: 21–30; 3: ≥31), cigars (0: ≤2; 1: 3–5; 2: 6–7; 3: ≥8), cigarillos (0: ≤5; 1: 6–10; 2: 11–15; 3: ≥16), and pipes (0: ≤4; 1: 5–8; 2: 9–12; 3: ≥13).

Total FTND scores range from 0 to 10, with higher scores indicating greater dependence. Participants with complete data received raw sum scores. For those missing 1–2 items, scores were prorated using personal weighted imputation: the observed sum was divided by the maximum possible score for the completed items, then rescaled to the full 10-point range.${FTND}_{prorated}=\left( \frac{Observed sum of answered items}{Maximum possible for those items} \right) \times10$. This approach accounts for the differing response ranges across items (i.e., Items 1 and 4 are scored 0–3, while Items 2, 3, 5, and 6 are scored 0–1). Participants missing three or more items were excluded from FTND analyses.

##### *Alcohol use disorder*

In Lifelines, lifetime AUD was assessed at Wave 3 (2019–2023) using the 11 DSM-5 diagnostic criteria [3] among participants who reported ever drinking alcohol, administered via self-report digital questionnaire. Participants responded retrospectively, reflecting on the 12-month period when their drinking was at its heaviest. The criteria assessed were:

1) drinking in larger amounts or for longer than intended
2) persistent desire or unsuccessful efforts to cut down or control use
3) substantial time spent obtaining, using, or recovering from alcohol
4) craving or strong urge to drink
5) failure to fulfill major role obligations
6) continued use despite social or interpersonal problems
7) giving up or reducing important activities because of drinking
8) recurrent use in physically hazardous situations
9) continued use despite physical or psychological harm
10) tolerance;
11) withdrawal (presence of withdrawal symptoms and/or drinking to relieve or avoid withdrawal).

Each criterion was coded as present (1) or absent (0); criterion 11 was coded as present if either withdrawal sub-item was endorsed.

To handle missing data while preserving diagnostic validity, we applied a missing-tolerant scoring strategy yielding both a continuous estimated symptom count and a categorical diagnosis of AUD assigned with explicit certainty rules, adapted from the PhenX Toolkit of the National Epidemiologic Survey on Alcohol and Related Conditions III [4] (NESARC-III, <https://www.phenxtoolkit.org/toolkit_content/supplemental_info/atos/additional_info/NESARC_III_Manual.pdf>).

*Continuous symptom count.* For participants with complete data, the symptom count was the raw sum of endorsed criteria (range: 0-11). For participants missing 1-3 items, symptom counts were estimated using personal mean imputation: $AUD symptom count=Observed sum + (Personal mean\times Number of missing items).$ Participants missing more than 3 items were assigned a missing symptom count (NA) and excluded from further AUD classification.

*AUD diagnosis.* Lifetime AUD was defined as the endorsement of at least 2 DSM-5 criteria. Participants were classified as cases (1) if they had endorsed at least 2 criteria, as the diagnostic threshold was met with certainty regardless of any missing items. They were classified as non-cases (0) only when the threshold could not be reached even if all missing items had been endorsed (i.e., endorsed 0 criteria with at most 1 missing item, or 1 criterion with no missing items). All remaining participants were assigned missing diagnostic status (NA), as the available data were insufficient to determine whether the 2-symptom threshold had been met.

*Severity classification.* Following DSM-5 guidelines, AUD severity was classified as mild (2-3 symptoms), moderate (4-5 symptoms), or severe (≥6 symptoms), using the same certainty-based approach. Mild AUD was assigned for 2 endorsed criteria with at most 1 missing item, or 3 endorsed criteria with no missing items. Moderate AUD was assigned for 4 endorsed criteria with at most 1 missing item, or 5 endorsed criteria with no missing items. Severe AUD was assigned for 6 or more endorsed criteria regardless of missingness. Severity was otherwise coded as missing when missing data could plausibly shift classification across a severity boundary.

In the BLTS (via the 19Up study) [5], AUD was assessed using the same 11 DSM-5 criteria [3]. Data were collected between 2009 and 2016. AUD diagnosis was based solely on available DSM-5 items: all participants had at least 7 items observed, and most (59%) had complete data on all 11 items. Participants were classified as having AUD if at least 2 observed criteria were endorsed. No mean imputation of missing items was applied, and participants were retained in analyses regardless of missing AUD items.

##### *Quality control and imputation of genotype data*

Lifelines participants were genotyped using three different arrays: the Illumina CytoSNP-12v2 array, the Infinium Global Screening Array® (GSA) MultiEthnic Disease Version 1.0, and the FinnGen Thermo Fisher Axiom® custom array, in order of release. Quality control (QC) of marker and samples were performed separately per array. For the CytoSNP array (released in 2020), quality control (QC) involved filtering SNPs with a minor allele frequency (MAF) above 0.001, a Hardy-Weinberg equilibrium (HWE) p-value >1e-4. A call rate threshold of 0.95 was used for both markers and samples. Sample QC included principal component analysis (PCA) to detect population outliers, and removal of duplicates, individuals with high heterozygosity and ambiguous sex, resulting in 249,249 markers and 15,422 samples. The UMCG Genetics Lifelines Initiative (UGLI) release 1 underwent a two-step QC for marker and sample missingness thresholds (from <80% to <99%), removing monomorphic markers (MAF = 0) and those with HWE p-value >1e-6. Samples with heterozygosity >4 standard deviations from the mean, duplicates, and ambiguous sex were excluded, yielding 548,029 markers and 36,339 samples. The UGLI release 2 (Affymetrix array) followed a similar two-step QC for call rates (first <80%, then <99%), removing markers with HWE p-value >1e-10 and MAF <0.02, and excluding samples with heterozygosity >4 SD from the mean, duplicates, and those with sex or family discrepancies, resulting in 462,731 markers and 28,249 samples. All arrays were imputed using the Haplotype Reference Consortium (HRC) panel (<http://www.haplotype-reference-consortium.org>) via the Sanger Imputation Service (http://imputation. sanger.ac.uk). Population stratification was examined using PCA with samples from the 1000 Genomes Project, retaining only individuals of European ancestry. Post-imputation, we filtered for imputation quality (INFO >0.8) and MAF >0.05 and selected high-quality markers (HapMap3+) in each array. We then selected markers that were available in all three arrays, resulting in 1,161,061 common markers. This overlap allowed for reliable haplotype comparison across parent-offspring pairs in the sample of 19,235 offspring with at least one genotyped parent, before matching with phenotype data. Detailed QC reports for each array are available on the Lifelines wiki (<http://wiki-lifelines.web.rug.nl/>).

##### *Non-transmitted alleles inference*

We applied HINTA (Haplotype-based Inference of Non-Transmitted Alleles), a validated approach for distinguishing transmitted and non-transmitted alleles in parent-offspring pairs and trios[1]. By including parent-offspring pairs and not just trios, this approach improves sample size, statistical power and generalizability.

Briefly, we used SHAPEIT5 to estimate haplotypes including pedigree information [6]. Offspring haplotypes were then compared to parental haplotypes using tiles of 150 adjacent markers on each chromosome. The best match between the parent and offspring tiles, taking recombination spots into account, was used to determine which parental tiles were transmitted to the offspring. The remaining non-transmitted alleles were recorded in a separate dataset, and for parent-offspring pairs, the non-transmitted alleles of the parent who was not genotyped were set as missing. This method was validated by comparison with standard software in parent-offspring trios and found a concordance rate for the non-transmitted alleles of 99.8%. Furthermore, the identification of non-transmitted alleles was confirmed to be unaffected by missing parental data through simulations of pairs from trios [1]. The same procedure was applied to both Lifelines and BLTS.

#### References

1.	Trindade Pons, V., et al., *Using parent-offspring pairs and trios to estimate indirect genetic effects in education.* Genetic Epidemiology, 2024. **48**(4): p. 190–199.

2.	Heatherton, T.F., et al., *The Fagerström test for nicotine dependence: a revision of the Fagerstrom Tolerance Questionnaire.* British journal of addiction, 1991. **86**(9): p. 1119–1127.

3.	Association, A.P., *Diagnostic and statistical manual of mental disorders: DSM-5™, 5th ed*. Diagnostic and statistical manual of mental disorders: DSM-5™, 5th ed. 2013, Arlington, VA, US: American Psychiatric Publishing, Inc. xliv, 947–xliv, 947.

4.	Toolkit, P., *The Alcohol Use Disorder and Associated Disabilities Interview Schedule—Diagnostic and Statistical Manual of Mental Disorders—Fifth Edition Version (AUDADIS–5) Alcohol and Drug Use Disorders Scoring Algorithms.*

5.	Gillespie, N.A., et al., *The Brisbane Longitudinal Twin Study: Pathways to Cannabis Use, Abuse, and Dependence project-current status, preliminary results, and future directions.* Twin Res Hum Genet, 2013. **16**(1): p. 21–33.

6.	Hofmeister, R.J., et al., *Accurate rare variant phasing of whole-genome and whole-exome sequencing data in the UK Biobank.* Nature Genetics, 2023. **55**(7): p. 1243–1249.

### Supplementary Tables and Figures

| Supplementary Table 1. Bivariate correlations among AUD and ND outcomes in the full Lifelines sample | | | | | |
| --- | --- | --- | --- | --- | --- |
|  | FTND_sum_ | CPD | AUD diagnosis | AUDsx | MaxDrinks |
| FTND_sum_ | 1 |  |  |  |  |
| CPD | **.63** | 1 |  |  |  |
| AUD diagnosis | **.20** | **.15** | 1 |  |  |
| AUDsx | **.17** | **.13** | **.82** | 1 |  |
| MaxDrinks | **.20** | **.18** | **.41** | **.36** | 1 |
| Note. FTND_sum_ = Fagerström Test for Nicotine Dependence total score; CPD = cigarettes per day; AUD diagnosis = lifetime DSM-5 AUD diagnosis; AUDsx = AUD symptom count; MaxDrinks = maximum alcoholic drinks in 24 hours. Pearson correlations were used for continuous variable pairs and point-biserial correlations for binary-continuous pairs. This table is based on the full Lifelines sample (*N*s ranged from 20,210 to 25,535). All reported correlations are significant at *p* < .001. | | | | | |

| Supplementary Table 2. Bivariate correlations between paternal and maternal transmitted and non-transmitted PGS in the Lifelines genotyped family sample | | | | | | | | | | | | |
| --- | --- | --- | --- | --- | --- | --- | --- | --- | --- | --- | --- | --- |
|  | (1) | (2) | (3) | (4) | (5) | (6) | (7) | (8) | (9) | (10) | (11) | (12) |
| (1) PGS_TUD_T_p_ | 1 | .017^*^ | .036^***^ | .014 |  |  |  |  |  |  |  |  |
| (2) PGS_TUD_T_m_ |  | 1 | .02 | .003 |  |  |  |  |  |  |  |  |
| (3) PGS_TUD_NT_p_ |  |  | 1 | .057^**^ |  |  |  |  |  |  |  |  |
| (4) PGS_TUD_NT_m_ |  |  |  | 1 |  |  |  |  |  |  |  |  |
| (5) PGS_PAU_T_p_ | .305^***^ | .002 | .022^*^ | -.008 | 1 | .008 | -.007 | .009 |  |  |  |  |
| (6) PGS_PAU_T_m_ | .023^**^ | .310^***^ | .020 | .007 |  | 1 | .014 | .013 |  |  |  |  |
| (7) PGS_PAU_NT_p_ | .005 | .002 | .311^***^ | .022 |  |  | 1 | .049^**^ |  |  |  |  |
| (8) PGS_PAU_NT_m_ | .012 | .015 | .026 | .290^***^ |  |  |  | 1 |  |  |  |  |
| (9) PGS_SUD_T_p_ | .349^***^ | .006 | .027^**^ | -.002 | .626^***^ | .012 | .004 | .005 | 1 | .015^*^ | .024^*^ | -.004 |
| (10) PGS_SUD_T_m_ | .015^*^ | .345^***^ | .008 | .006 | .017^*^ | .633^***^ | .008 | .010 |  | 1 | .000 | .022^*^ |
| (11) PGS_SUD_NT_p_ | .003 | .011 | .364^***^ | .013 | .015 | .000 | .637^***^ | .029 |  |  | 1 | .026 |
| (12) PGS_SUD_NT_m_ | .018^*^ | .006 | .028 | .344^***^ | -.003 | .011 | .012 | .632^***^ |  |  |  | 1 |
| Note. Sample sizes for transmitted and non-transmitted polygenic scores (PGS) in offspring ranged from 3,267 (mother-father-offspring trios sample) to 19,233 (trios and parent-offspring pairs sample). Pearson correlations were used for continuous variables. Significance levels: ^∗^*p* < .05; ^∗∗^*p* < .01; ^∗∗∗^*p* < .001. TUD = tobacco use disorder; PAU = problematic alcohol use; SUD = substance use disorder (general addiction liability); T= transmitted; NT = non-transmitted; p = paternal; m = maternal. | | | | | | | | | | | | |

| Supplementary Table 3. Meta-analysis of transmitted and non-transmitted polygenic score effects on alcohol use disorder (AUD) across Lifelines and BLTS | | | | | | | | | |
| --- | --- | --- | --- | --- | --- | --- | --- | --- | --- |
|  | | | **PGS_T_** | | |  | **PGS_NT_** | | |
| **Predictor** | **Outcome** | **N** | **β_T_ (SE) / OR** | **95% CI** | **p** |  | **β_NT_ (SE) / OR** | **95% CI** | **p** |
| **PGS_PAU_** | | | | | | | | | |
| Lifelines | | | | | | | | | |
|  | *AUDsx* | 4,862 | .078 (.014) | .050, .105 | <.001 |  | -.007 (.014) | -.035, .020 | .598 |
|  | *AUD diagnosis* | 4,861 | 1.196 | 1.105, 1.294 | <.001 |  | 0.959 | .888, 1.035 | .277 |
| BLTS | | | | | | | | | |
|  | *AUDsx* | 1,368 | .126 (.027) | .072, .180 | <.001 |  | -.030 (.028) | -.084, .024 | .280 |
|  | *AUD diagnosis* | 1,368 | 1.319 | 1.145, 1.520 | <.001 |  | .894 | .777, 1.029 | .119 |
| Meta-analysis | | | | | | | | | |
|  | *AUDsx* | 6230 | .096 (.023) | .051, .142 | <.001 |  | -.012 (.013) | -.036, .013 | .354 |
|  | *AUD diagnosis* | 6229 | 1.233 | 1.128, 1.348 | <.001 |  | .944 | .882, 1.009 | .091 |
| **PGS_SUD_** | | | | | | | | | |
| Lifelines | | | | | | | | | |
|  | *AUDsx* | 4,862 | .061 (.014) | .033, .089 | <.001 |  | -.014 (.014) | -.042, .014 | .322 |
|  | *AUD diagnosis* | 4,861 | 1.113 | 1.031, 1.202 | .006 |  | .975 | .903, 1.053 | .520 |
| BLTS | | | | | | | | | |
|  | *AUDsx* | 1,368 | .117 (.028) | .063, .172 | <.001 |  | -.033 (.028) | -.087, .022 | .242 |
|  | *AUD diagnosis* | 1,368 | 1.295 | 1.122, 1.494 | .001 |  | .869 | .755, 1.000 | .051 |
| Meta-analysis | | | | | | | | | |
|  | *AUDsx* | 6230 | .084 (.028) | .030, .138 | .002 |  | -.018 (.013) | -.042, .007 | .155 |
|  | *AUD diagnosis* | 6229 | 1.186 | 1.024, 1.372 | .023 |  | .935 | .839, 1.042 | .225 |
| Note. AUDsx = AUD symptom count; *β* = standardized regression coefficient (for continuous AUD symptom count); OR = odds ratio (for binary AUD diagnosis); SE = standard error; PGS_PAU_= polygenic score for problematic alcohol use; PGS_SUD_ = polygenic score for substance use disorders. PGS_T_= transmitted PGS; PGS_NT_ = non-transmitted PGS. BLTS = Brisbane Longitudinal Twin Study. Meta-analysis used fixed-effects inverse-variance weighting across cohorts. | | | | | | | | | |

Supplementary Table 4. Mediation analysis: transmitted and non-transmitted polygenic scores for tobacco use disorder on offspring cigarettes per day

|  | Model A (Smoking) β [95% CI] | Model B (Education) β [95% CI] | Model C (Income) β [95% CI] | Model D (Joint Parallel) β [95% CI] |
| --- | --- | --- | --- | --- |
| **PGS_T_TUD_ (Mother)** | | | | |
| Total effect | .081 [.056, .106] ^*^ | .078 [.053, .104] ^*^ | .078 [.053, .104] ^*^ | .081 [.054, .106] ^*^ |
| Direct effect | .059 [.033, .085] ^*^ | .076 [.050, .102] ^*^ | .077 [.052, .103] ^*^ | .057 [.029, .083] ^*^ |
| Mediated effect |  |  |  |  |
| *Via smoking* | .022 [.015, .030] ^*^ | — | — | .022 [.015, .030] ^*^ |
| *Via education* | — | .002 [.000, .005] ^*^ | — | .002 [.000, .004] |
| *Via income* | — | — | .000 [-.001, .003] | — |
| **PGS_T_TUD_ (Father)** | | | | |
| Total effect | .093 [.070, .118] ^*^ | .094 [.070, .119] ^*^ | .095 [.070, .120] ^*^ | .093 [.070, .117] ^*^ |
| Direct effect | .089 [.067, .115] ^*^ | .093 [.069, .118] ^*^ | .095 [.070, .120] ^*^ | .088 [.065, .112] ^*^ |
| Mediated effect |  |  |  |  |
| *Via smoking* | .004 [.001, .009] ^*^ | — | — | .004 [.001, .009] ^*^ |
| *Via education* | — | .001 [-.001, .003] | — | .001 [-.002, .003] |
| *Via income* | — | — | .000 [-.001, .001] | — |
| **PGS_T_TUD_: Maternal minus paternal mediated effect** | | | | |
| M-F: Via smoking | .018 [.009, .027] ^*^ | — | — | .018 [.009, .027] ^*^ |
| M-F: Via education | — | .001 [-.002, .005] | — | .001 [-.002, .005] |
| M-F: Via income | — | — | .000 [-.002, .004] | — |
| **PGS_NT_TUD_ (Mother)** | | | | |
| Total effect | .015 [-.016, .047] | .017 [-.015, .048] | .015 [-.015, .047] | .017 [-.014, .049] |
| Direct effect | .002 [-.030, .033] | .014 [-.019, .045] | .016 [-.015, .046] | .000 [-.029, .031] |
| Mediated effect |  |  |  |  |
| *Via smoking* | .014 [.007, .021] ^*^ | — | — | .014 [.007, .021] ^*^ |
| *Via education* | — | .004 [.001, .007] ^*^ | — | .003 [.000, .006] |
| *Via income* | — | — | .000 [-.001, .001] | — |
| **PGS_NT_TUD_ (Father)** | | | | |
| Total effect | .043 [.005, .082] ^*^ | .047 [.007, .083] ^*^ | .047 [.006, .083] ^*^ | .044 [.006, .081] ^*^ |
| Direct effect | .037 [-.001, .074] | .045 [.005, .081] ^*^ | .047 [.006, .083] ^*^ | .036 [-.002, .074] |
| Mediated effect |  |  |  |  |
| *Via smoking* | .007 [.002, .011] ^*^ | — | — | .007 [.002, .012] ^*^ |
| *Via education* | — | .001 [-.001, .004] | — | .001 [-.002, .003] |
| *Via income* | — | — | .000 [-.002, .002] | — |
| **PGS_NT_TUD_: Maternal minus paternal mediated effect** | | | | |
| M-F: Via smoking | .007 [-.002, .016] | — | — | .007 [-.002, .016] |
| M-F: Via education | — | .003 [-.002, .008] | — | .003 [-.002, .008] |
| M-F: Via income | — | — | .000 [-.003, .002] | — |
| Note. Model A = parental smoking as single mediator; Model B = parental education as single mediator; Model C = household income as single mediator; Model D = parental smoking and education as parallel mediators (income excluded based on null single-mediator results). All estimates are standardized (β). Models were estimated using full information maximum likelihood (FIML) with cluster-robust standard errors, adjusting for offspring sex and age. The total analytic sample passed to each model was *N* = 19,233. Observed *N*s varied across variables due to the data availability: transmitted PGS *N* = 19,233 (both parents); non-transmitted PGS *N* = 13,236 (maternal) and 9,264 (paternal); parental CPD *N* = 6,472 (maternal) and 5,152 (paternal); parental education *N* = 12,965 (maternal) and 9,095 (paternal); parental income *N* = 10,114 (maternal) and 7,800 (paternal); offspring CPD *N* = 5,972. Mediated effects and 95% CIs estimated via Monte Carlo simulation (2,000 replications). * = 95% CI excludes zero. — = pathway not included in model. PGS_T_ = transmitted polygenic score; PGS_NT_ = non-transmitted polygenic score; TUD = tobacco use disorder; CPD = cigarettes per day. | | | | |

Supplementary Figure 1. Forest plots of transmitted and non-transmitted polygenic score associations with alcohol use disorder outcomes across Lifelines and BLTS cohorts

Note. Meta-analysis of transmitted (PGS_T_) and non-transmitted (PGS_NT_) polygenic score associations with AUD symptom count and AUD diagnosis, using problematic alcohol use (PGS_PAU_; left panels) and substance use disorders (PGS_SUD_; right panels) scores. Individual study estimates are shown with 95% confidence intervals (dashed lines). Pooled estimates from random-effects (RE) meta-analysis are represented by diamonds, with blue indicating statistically significant effects and red indicating non-significant effects. Continuous outcomes (AUD symptom count) are shown as standardized *β* coefficients; binary outcomes (AUD diagnosis) are shown as odds ratios on a log scale. Total meta-analytic sample: *N* = 6229 (Lifelines *N* = 4861; BLTS *N* = 1,368). AUD = alcohol use disorder; BLTS = Brisbane Longitudinal Twin Study.
